# The SENSOR System: Using Standardized Data Entry and Dashboards for Review of Scientific Studies Using the Clinical Applications of Psychedelics as an Illustration

**DOI:** 10.1101/2022.01.14.22269304

**Authors:** Latifah Kamal, Major Pauline Godsell, Bryce Mulligan, Stefan Eberspaecher, Danny Myint, LCol Markus Besemann, Amir Minerbi, Gaurav Gupta

## Abstract

**Introduction:** Literature reviews are useful tools for communicating the breadth of scientific discovery for a given topic. Irrespective of the nature of the review, data should be communicated in effective, easy to understand ways. In trying to address these limitations of traditional scientific reviews, we propose using dynamic data driven displays that have been used in multiple other industries to improve communication and decision making. Given the recent interest in the clinical applications of psychedelics for various mental health issues, we chose to test the SENSOR System (Standardized Data Entry and Dashboards for Review of Scientific Studies) as an alternative for an existing review article.

**Methods:** To validate the SENSOR System, an existing review with a topical, heterogenous, and growing set of studies was selected. In this case we chose the Wheeler et al. review on psychedelics in clinical practice where articles had already been preselected and reviewed. Detailed discussion of this review and the cited papers preceded designing the content and shared links for a Google Form for data intake, Google Drive for article access, and Google Sheets linked to the form intake data.

**Results:** A total of 46 study entries were made by 2 team members, including 3 articles published since the review to demonstrate the ease of updating the system Various representations of the Google Forms intake data in the SENSOR System dashboard are presented.

**Discussion:** Visual representation of review studies using a dashboard proved feasible and advantageous for numerous reasons. As the technology and guidelines for these systems evolve there is an opportunity to standardize reporting, centralize legacy datasets, streamline the submission process, improve collaboration between researchers, measure relative contribution of participating authors, and improve patient involvement. For the use case of clinical applications of psychedelics, limitations of conveying data accurately includes heterogeneity of study design, dosing, indications, and outcome measures.

**Conclusion:** Creation of a system for standardized data entry and dashboards for reviews of scientific studies is a feasible alternative and/or adjunct to the dissemination of summaries through traditional scientific review. There are numerous proposed advantages of the flexible, dynamic, and graphical display that requires further validation.

## Introduction

Literature reviews are useful tools for communicating the breadth of scientific discovery for a given topic. Given the ever-increasing output of scientific publications, synthesis of this data is becoming necessary for summaries of this research. [1,2,3] These reviews allow for more efficient understanding of a body of study, hypothesis generation and understanding of the limits of clinical applications.

Various types of reviews have evolved over time. Format variations include scale (mini vs full), expression (descriptive vs. integrative) and type (narrative vs. systematic) reviews. [4,5,6] Therefore, the process of writing reviews can be complex, depending on the nature of the review, the intended goal, and the required structure. [7] In general, these steps include topic selection, literature search, critical appraisal, synthesis, report writing, and revision based on feedback. [8,9] The quality of the work product is predicated on systematically reducing the impact of bias and where possible, combining data to augment the power of the analysis (i.e. meta-analysis). [10,11]

Irrespective of the nature of the review, data should be communicated effectively, in an easy to understand way. Images are better for comprehension than numbers or words, suggesting graphics and pictures could facilitate synthesis of complex scientific communication. [12,13,14,15,16] Considerable literature has highlighted strategies for enhancing visual communications in scientific publications. These include a focus on simplicity, object/attribute encoding type and colors, visual patterning and distinctions, suitable axis selection, and carefully considered data axis, transformation, aggregation, and time series sequential. [15] Nevertheless even with these guidelines, the nature of scientific reviews has been largely static overtime. These communications can be time consuming to write, vast amounts of information can be difficult to synthesize, relative comparisons of data can be challenging to make depending on the data and in some cases outdated by the time of publication.

In trying to address these limitations of traditional scientific reviews, we propose using dynamic data driven visual displays that have been used in multiple other industries to improve communication and decision-making. These dashboards theoretically can also exploit perceptual capacities to enhance cognition, but should be tailored to the use, case and/or user. [17] The dashboards themselves can contain both tabular and graphical information that can be manipulated by data filters and slicers.

We hypothesize that an appropriate data input system and spreadsheet software could facilitate development of a dashboard as a standalone and/or complementary tool for a traditional scientific review. Given the recent interest in the clinical applications of psychedelics for various mental health issues, we chose to test the SENSOR System (Standardized Data Entry and Dashboards for Review of Scientific Studies) as an alternative for an existing review article written by Wheeler et al. [18]

## Methods

To validate the SENSOR System, an existing review with a topical, heterogenous, and growing set of studies were selected. [18] In this case we chose the Wheeler et al. review on psychedelics in clinical practice where articles had already been preselected and reviewed. Each participating author was assigned their own articles, but any concerns regarding data input were discussed with the appropriate team member. Detailed discussions of the review and the cited papers preceded designing shared links for a Google Form for data intake, Google Drive for article access, and Google Sheets linked to the form intake data.

The Google Form questions included a combination of short and long answer data, multiple checkboxes, and multiple choices. Table 1 shows how variables were assigned to questions subtypes, and “other” options were included, where possible. While flexible, questions were ordered in a way to facilitate data entry. For numerical inputs, care was taken to standardize input (e.g. time, absolute percentage of improvement for a given outcome measure, etc.). Where responder rate was reported, the total trial size was separately recorded, which allowed for bubble graphs to be generated for the dashboard. For medications, an average dose was calculated, where amounts between dosing sessions differed and were normalized for use in a 70kg participant. Patients were classified by their main and secondary indications where necessary, for instance those with anxiety and depression in the case of palliative conditions or addiction to various substances such as smoking or alcohol.

**Table 1:** Google Form Variables and Questions Subtype assignment.

| Short Answer | Long Answer | Checkboxes | Multiple Choice |
| --- | --- | --- | --- |
| Email | Comments | Study Country(ies) | Indication(s) |
| Date |  | Type of Study |  |
| Year of Publication |  | Nature of Controls |  |
| Author Name |  | Medication Used |  |
| G-Drive article link |  | Medication Analogs |  |
| Primary City |  | Administration Route |  |
| Trial Size (Controls and Patients) |  | Control Group Type |  |
| Average Dose |  | Outcome Measure |  |
| Average Control Dose |  | Interval |  |
| Percent response |  | Adverse Events |  |
| Trial Length |  | Outcome Measures |  |
|  |  | Limitations |  |

When considering response to treatment, allowance for multiple outcome measures were made in the one intake form, but functionally, only the primary outcome was recorded. Outcome measures were reported as percentages, based on the absolute improvement for a given scale (i.e. a change from 4-2 on a 10 point scale was a 20% improvement, not 50%) or responder rate if necessary (i.e. number of patients that achieved a certain percentage improvement).

The articles themselves were made available in a shared Google Drive and the links for each article were included in the form for ease of access in the future. The dashboard itself was created in Google Sheets with filters for publication year, medication, dose, location (i.e. city), percent improvement, author, indication and trial size. Graphs within the dashboard included bar graphs for trial size and percent response by both medication used and indication respectively. A bubble graph was also included, comparing percentage relief against dose with colors representing medication type and bubble size reflecting trial size. A map for study locations and pivot tables were incorporated to help with data presentation and to facilitate communication of information.

## Results

A total of 46 research paper entries were made by 2 team members, including 3 articles published since the review to demonstrate the ease of updating the system. This represents a total of 1237 patients, with an average response of 31%, spanning multiple continents, medications and indications. We note the presented data is impacted by the Wilkinson et al meta-analysis on ketamine for which there is no dosing data and the qualitative study by Swift et al for which no outcomes were presented. [19,20]

The data entered into the spreadsheet far exceeds what was included in the dashboard. See Figures 1-8 for full graphics. Below are the various representations of the Google Forms intake data in the SENSOR System dashboard. All aspects of the graphs are easily customizable using the slicers. The drop-down menus provide the opportunity to remove or add any data input immediately into the graphs. Currently, for ease of visibility, two medications (ibogaine and nitrous oxide) with high relative dosing regimens have been omitted from all represented data as seen in the slicers of figure 2. Given the entered data far exceeds what is included in the current dashboard (e.g. limitations, measurement tool, reviewer notes) multiple dashboards can be used in the future for a specific data set. Despite best efforts we concede not all calculations will be accurate, and encourage readers to focus on the benefits of the system itself.

**Figure 1:**
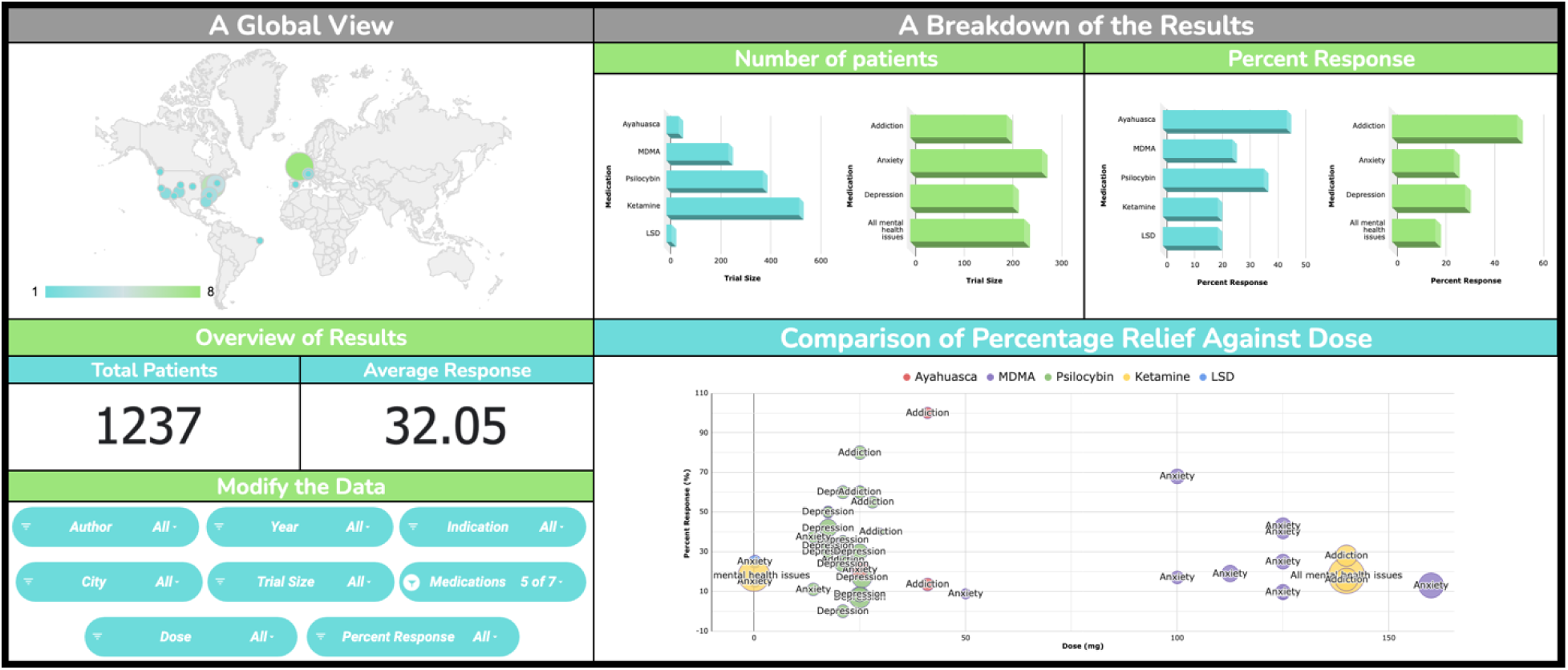
Global View of Dashboard.

**Figure 2:**
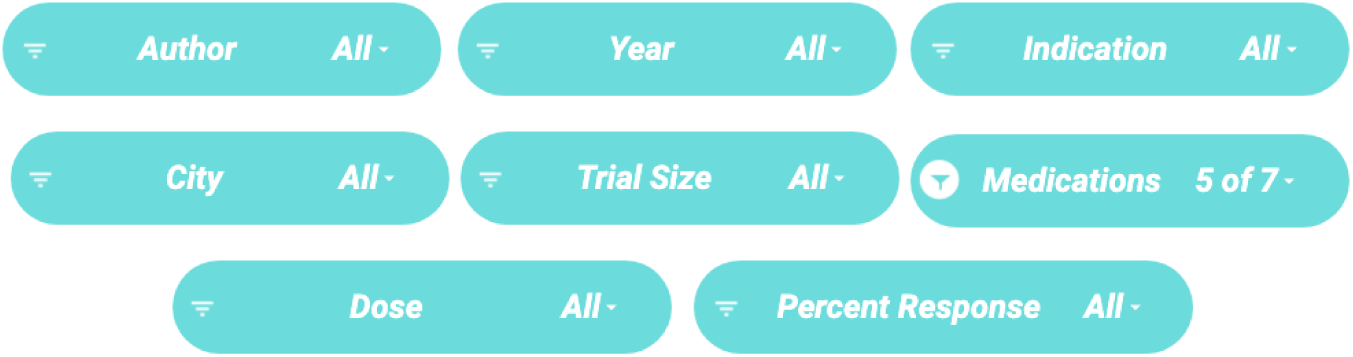
Slicers to Modify the Data.

**Figure 3:**
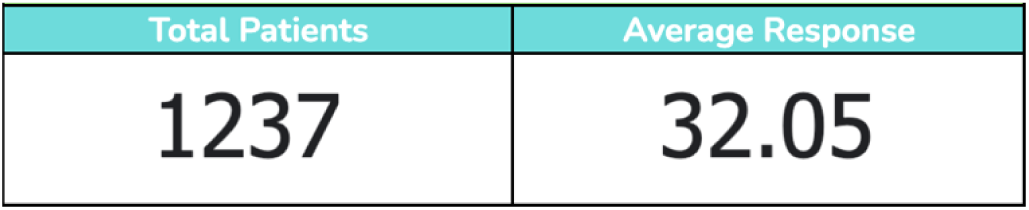
Overview of Results.

**Figure 4a:**
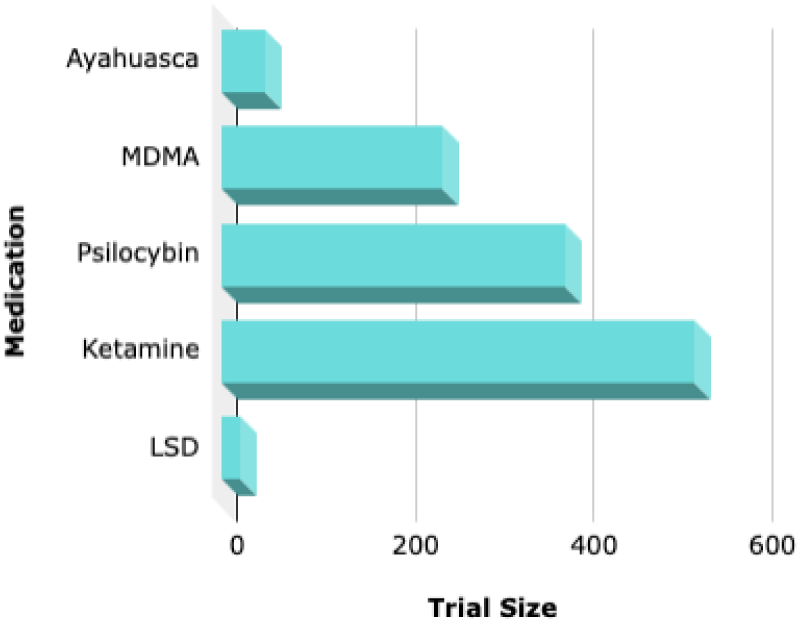
Trial Size by Medication Used.

**Figure 4b:**
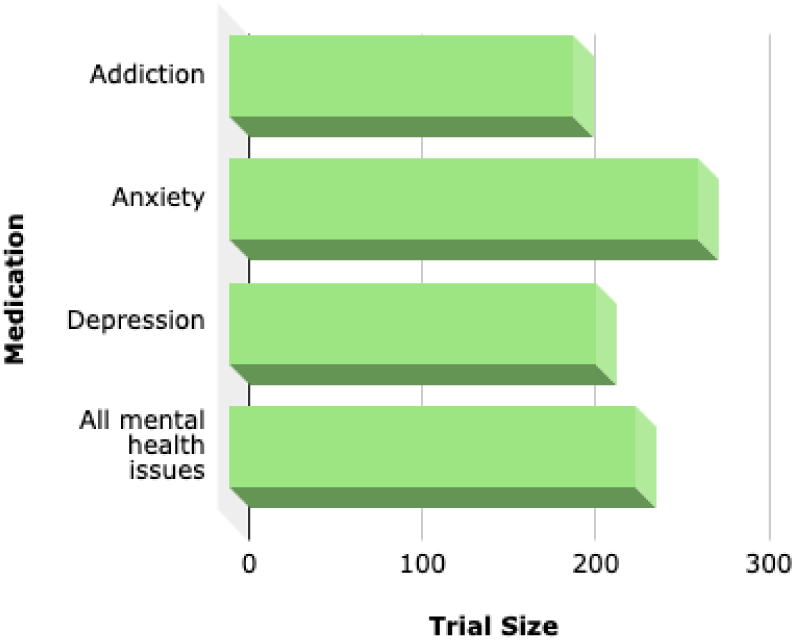
Trial Size by Indication.

**Figure 5a:**
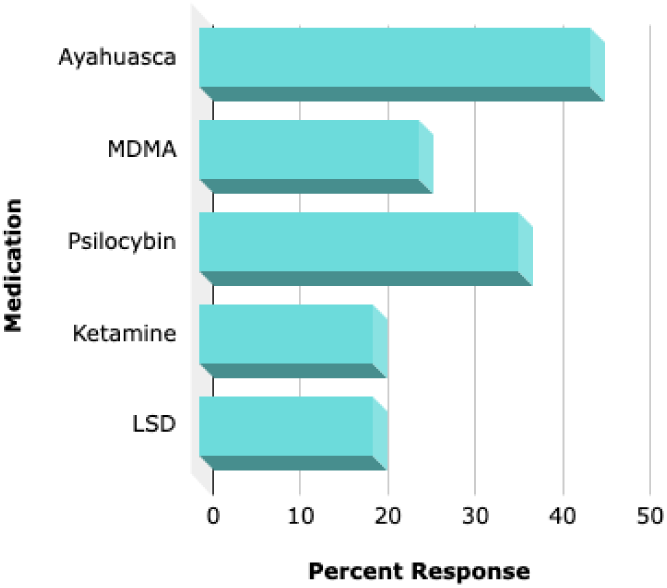
Percent Response by Medication Used.

**Figure 5b:**
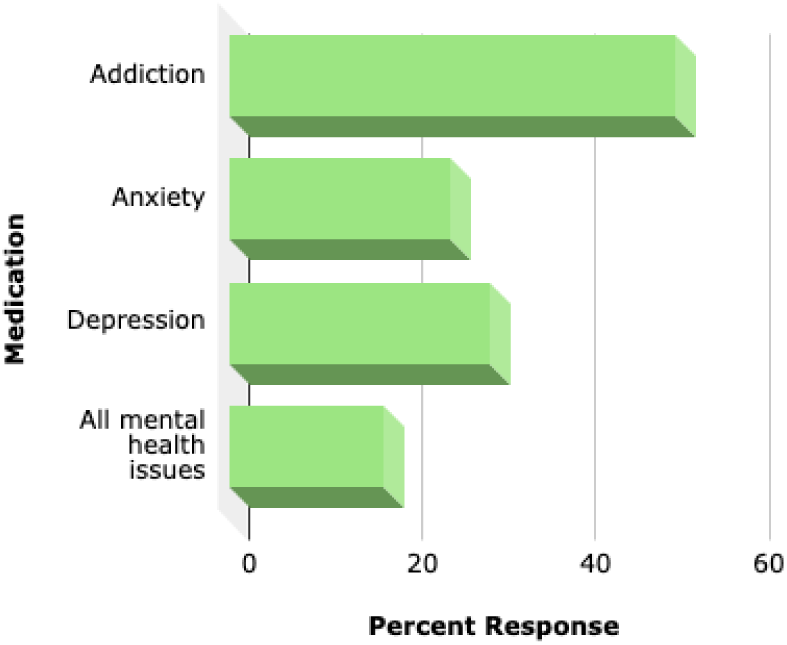
Percent Response by Indication.

**Figure 6:**
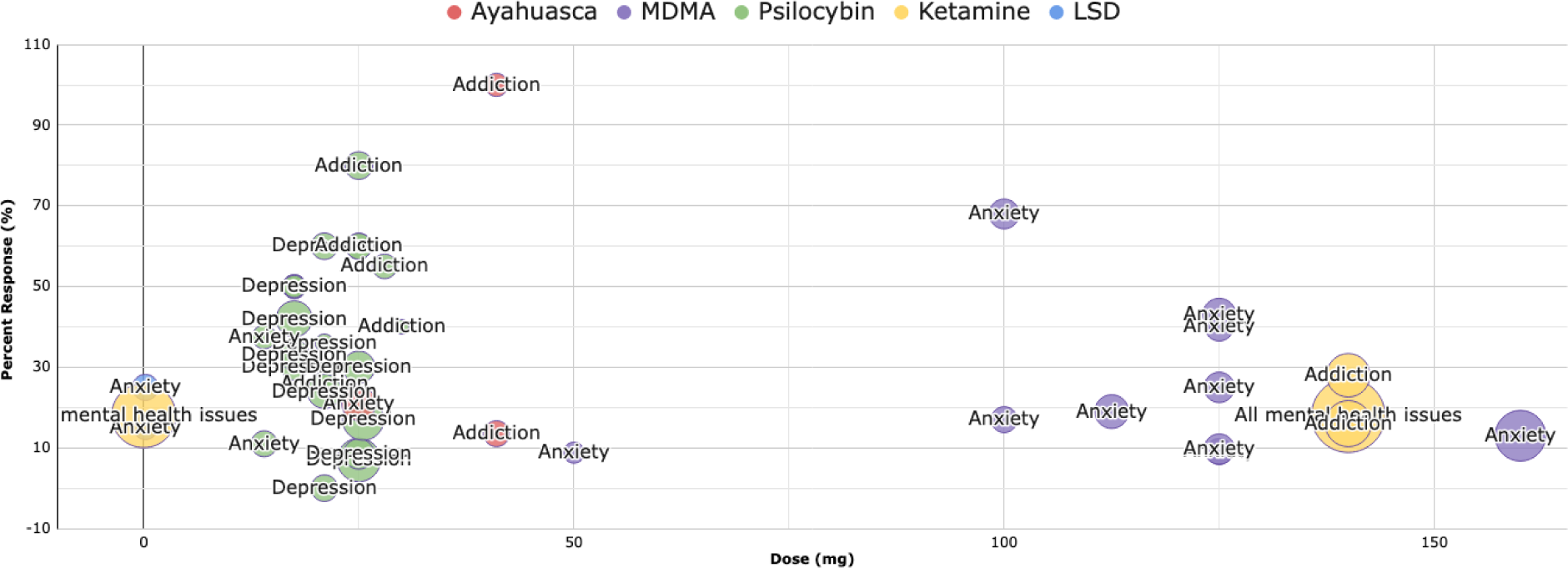
Comparison of Percentage Relief Against Dose.

**Figure 7:**
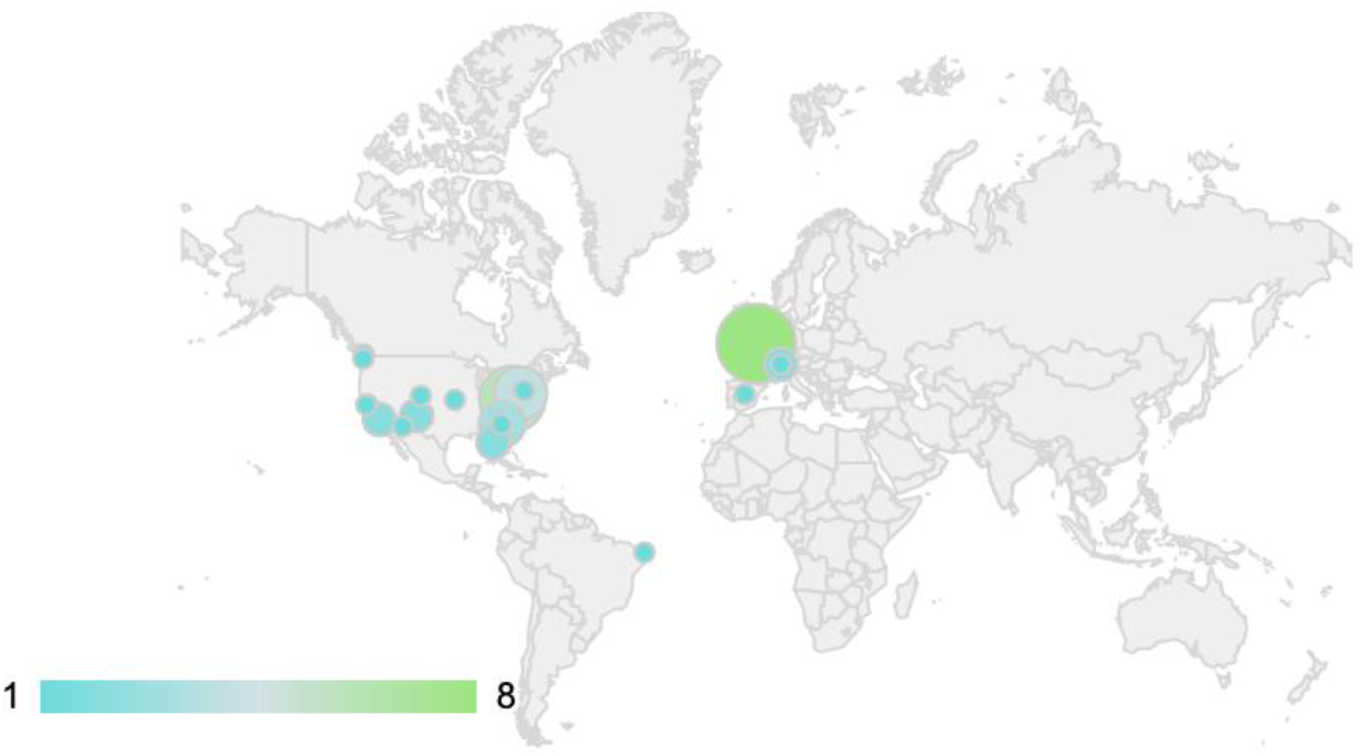
Map of Study Locations.

**Figure 8:**
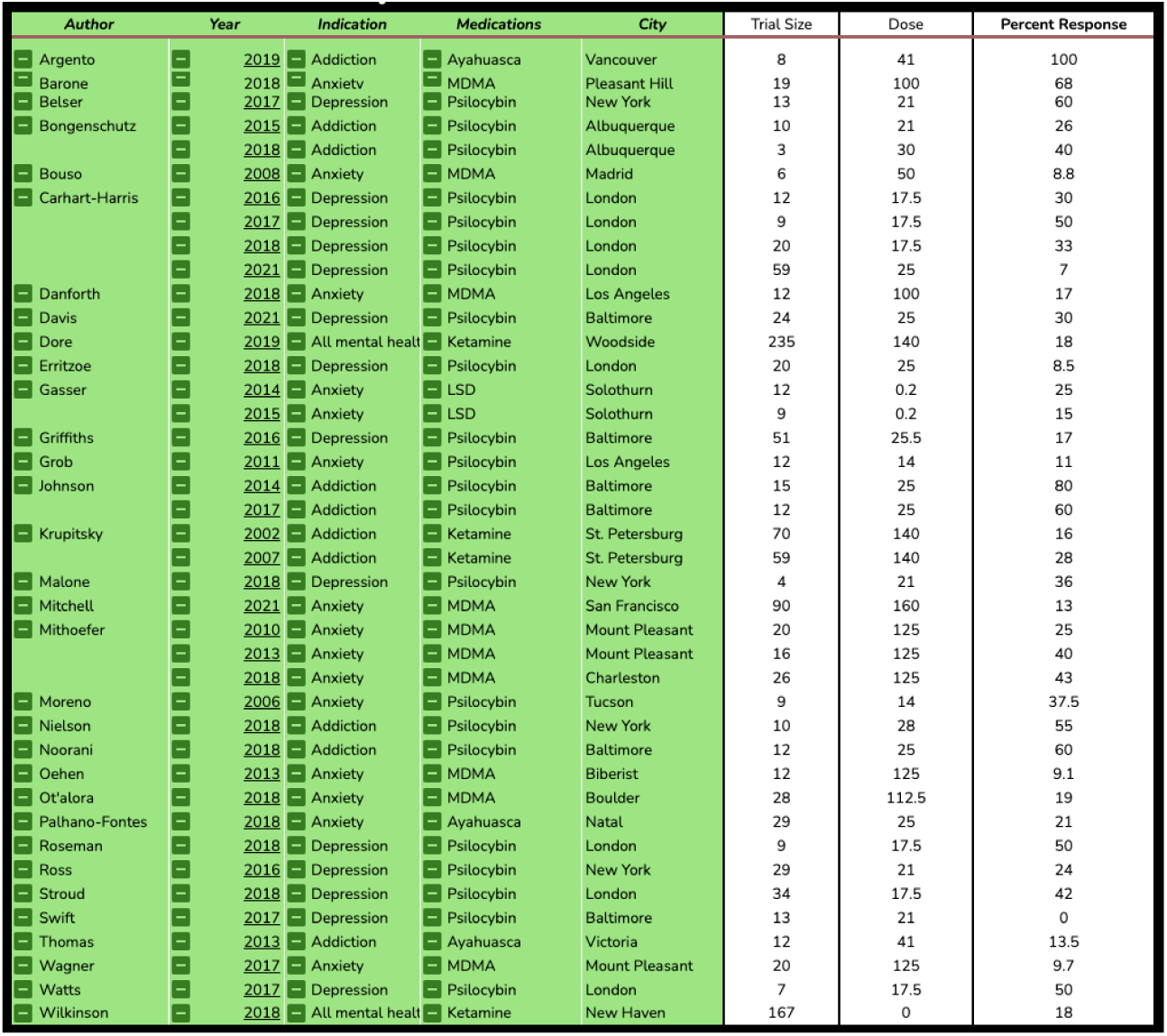
Pivot Tables.

## Discussion

The main goal of this study was to determine the feasibility of the using the SENSOR System for scientific reviews. The study specifically examined the technical process of data input and dashboard using an existing review article. Despite the inherent challenges of the literature examined, and the technological constraints, the data from the index trials were clearly and dynamically presented in the resulting dashboard.

We believe if employed properly this approach could offer an alternative to the traditional dissemination of scientific reviews in various scientific and clinical fields. The applications could include rebooting traditional review articles, journal or organization specific dashboards summarizing existing literature selectively open to certain authors, or systems designed to direct future inquiry for validation and discovery purposes. See table 3 for proposed advantages of the SENSOR System.

**Table 3:**
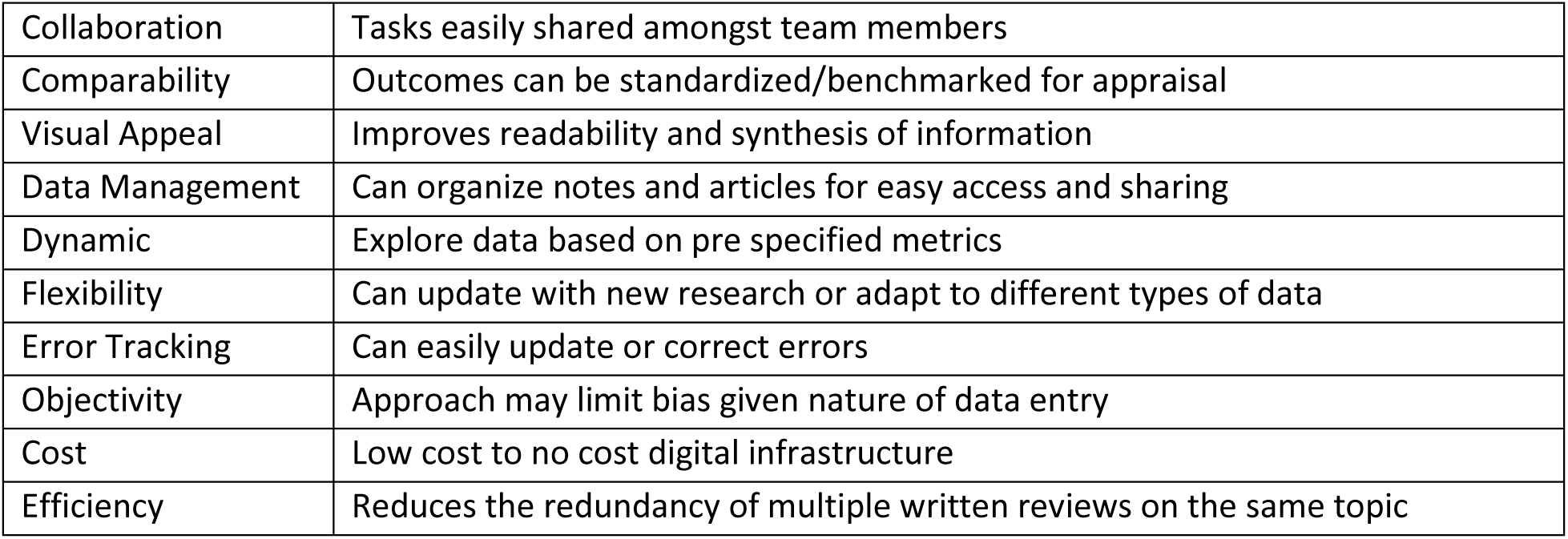
Proposed Advantages of the SENSOR System for Scientific Reviews.

As the technology and guidelines for these systems evolve there is an opportunity to standardize reporting, centralize legacy datasets, streamline the submission process, improve collaboration between researchers, measure relative contribution of participating authors, and improve patient involvement.

Limitations include accuracy of the conveyed data included heterogeneity of study design, dosing, indications, and outcome measures. In addition, further research would be required to formally test usability, applicability and adaptations when compared to the traditional non-digitized scientific review methods. Finally, organizations will have to ensure the appropriate infrastructure is in place to manage these systems overtime. This will require an understanding of the resources and effort for the various components such data entry, dashboard management and dissemination. For limitations and challenges see table 4.

**Table 4:** Challenges and Limitations of the SENSOR System in reviewing Clinical Applications of Psychedelics.

|  |  |
| --- | --- |
| Consistency | Determining whether to report responder rate vs absolute changes |
| Measures | Various heterogeneous outcomes measures used requiring standardized reporting |
| Methods | Various trial designs including qualitative and crossover which complicated interpretation and control comparisons |
| Results | Absolute changes compared to total outcome scored preferred where possible and total scoring available, whereas reports often focused on relative benefit |
| Completeness | Variable data types, methods of presentation and inclusiveness based on report therefore only results from primary scales were included |
| Critical Appraisal | Can be itemized to a degree, but insights and opinion for peer review can be limited depending on system design |
| Technology | Access and comfort with technology may limit utility for some authors |
| Overlap | Multiple studies from a single patient population were reported independently for this illustration but ideally treated as a single study with multiple outcomes in the future |
| Reporting | Not amenable static print or digital publications or traditional reporting structure |

## Conclusion

Creation of a system for standardized data entry and dashboards for reviews of scientific studies is a feasible alternative and/or adjunct to the dissemination of summaries through traditional scientific review. There are numerous proposed advantages of the flexible, dynamic, and graphical display that requires further validation.

## Data Availability

All data produced in the present work are contained in the manuscript

